# The effectiveness of preoperative oral carbohydrate loading on postoperative nausea and vomiting in adults receiving total intravenous anaesthesia compared to inhalational anaesthesia: A systematic review and meta-analysis

**DOI:** 10.1101/2025.06.16.25329650

**Authors:** Oya Gumuskaya, Hailey R Donnelly, Nick Glenn, Julee McDonagh, Anita Skaros, Sophie Liang, Brett G Mitchell, Luke Bendle, Sarah Aitken, Mitchell Sarkies

**Affiliations:** School of Health Sciences, Faculty of Medicine and Health, The University of Sydney, NSW, Australia; School of Nursing and Midwifery, Western Sydney University, Sydney, NSW, Australia; School of Health Sciences, College of Health, Medicine and Wellbeing, University of Newcastle, Newcastle, NSW, Australia; Food and Nutrition Research Program, Hunter Medical Research Institute, New Lambton Heights, NSW, Australia; Centre for Chronic & Complex Care Research, Blacktown Hospital, Western Sydney Local Health District, NSW, 2148 Australia; School of Nursing, Faculty of Science Medicine and Health, University of Wollongong NSW 2170 Australia; Department of Anaesthetics, Royal Prince Alfred Hospital, Sydney Local Health District, Camperdown, NSW, Australia; Department of Anaesthesia and Perioperative Medicine, Westmead Hospital, Western Sydney Local Health District, Westmead, NSW, Australia; Department of Anaesthesia and Pain Management, Concord Repatriation General Hospital, Sydney Local Health District, Camperdown, NSW, Australia; Sydney Medical School, Faculty of Medicine and Health, The University of Sydney, NSW, Australia; School of Nursing and Health, Avondale University, Lake Macquarie, NSW, Australia; Central Coast Local Health District, Gosford Hospital, Gosford, NSW, Australia; School of Nursing, Paramedicine and Midwifery, Australian Catholic University, Ballarat, Victoria, Australia; Concord Institute of Academic Surgery, Concord Repatriation General Hospital, Sydney Local Health District, Concord West, NSW, Australia; Implementation Science Academy, Sydney Health Partners, The University of Sydney, NSW, Australia

**Keywords:** Fasting, surgery, anaesthetics, emesis, PONV

## Abstract

**Background:** Preoperative oral carbohydrate loading is thought to reduce postoperative nausea and vomiting (PONV). However, it is unknown if the benefit of carbohydrate loading is maintained in the presence of total intravenous anaesthesia (TIVA). The aim of this systematic review was to determine whether oral carbohydrate loading reduced PONV compared to overnight fasting between adult elective surgery patients receiving TIVA or inhalational general anaesthesia.

**Methods:** A search of seven databases was conducted until March 2024. Randomised controlled trials conducted with patient aged 18 years or older were included. Two reviewers independently screened titles, abstracts and full texts, and assessed risk of bias using the Cochrane ROB-2 Tool. Study data was pooled using random effects meta-analyses.

**Results:** We included 26 studies in this review, and 25 in the meta-analyses (n=2,491). Preoperative oral carbohydrate loading reduced the overall risk (log risk ratio: –0.41, 95% CI –0.72 to –0.11, I2=30.92%) and severity (SMD: -0.46, 95% CI: -0.71 to -0.21, I2=66.59) of PONV, and pain severity (mean difference: -0.68, 95% CI: -1.13 to -0.22, I2=85.32%) compared to prolonged fasting when pooled across both anaesthesia approaches. The risk of PONV was reduced in patients receiving inhalational anaesthesia, but not TIVA, while the reduction in severity was more significant in TIVA.

**Conclusion:** Oral carbohydrate loading reduces the severity of PONV and pain, regardless of the anaesthesia approach, compared to prolonged fasting. These findings support the clinical advantages of oral carbohydrate loading for postoperative outcomes, regardless of anaesthesia approach.

## Introduction

Postoperative nausea and vomiting (PONV) affects approximately a third of general surgery patients. PONV is consistently described by patients as one of the most undesirable perioperative experiences[1–3]. Beyond the negative impact on patient experience, PONV is also associated with delayed recovery, prolonged hospital length of stay, and increased healthcare costs[4].

Prevention of PONV is a pillar of enhanced recovery pathways for various types of surgery to mitigate adverse effects, improve postoperative outcomes, and enhance patient satisfaction[1, 5]. Prolonged preoperative fasting exacerbates the metabolic stress response from surgery, contributing to increased insulin resistance, higher rates of surgical complications, and PONV [6–9]. Therefore, preoperative oral carbohydrate loading is suggested as a strategy to shorten preoperative fasting times, with evidence suggesting potential benefits for reducing metabolic disturbances and improving recovery [10, 11]. However, the evidence on the efficacy of preoperative oral carbohydrate loading in preventing PONV remains inconclusive and may contribute to why the adoption of reduced fasting practices is not more widespread [9, 12]. Multimodal strategies to address PONV, including total intravenous anaesthesia (TIVA) have been integrated into routine care with benefits demonstrated for patients at high risk of PONV [13, 14]. Despite this, prolonged fasting and PONV remain a common issue [15–19]. Additionally, most studies have not differentiated between patients receiving TIVA and those receiving inhalational anaesthesia, leaving a significant gap in understanding the interplay between anaesthetic techniques, carbohydrate loading, and PONV outcomes.

The primary aim of this systematic review was to evaluate the effects of preoperative oral carbohydrate loading compared to standard preoperative fasting on the risk and severity of PONV in adult patients undergoing elective surgery, and whether these effects differ between TIVA or inhalational general anaesthesia. The secondary aim was to evaluate the effects of preoperative oral carbohydrate loading on postoperative pain and hospital length of stay.

## Methods

This systematic review was registered with Prospero (CRD42021222171) and reported according to the Preferred Reporting Items for Systematic Review and Meta-Analyses statement (PRISMA)[20] .

### Search strategy

Seven electronic databases (MEDLINE, Science Direct, Scopus, CINAHL, Cochrane, Web of Science, and Clinical Key) were searched for peer-reviewed articles from inception until 1^st^ of March 2024. The search results were imported to a web-based reference management tool: Covidence^TM^ (Veritas Health Innovation, Melbourne, Australia). The reference lists of included articles were snowballed and duplicates removed. See Appendix 1 for the full search strategy.

### Selection criteria

Randomised controlled trials were included if they reported the effect of preoperative oral carbohydrate loading compared to overnight fasting on the incidence or severity of PONV, post-operative nausea, and post-operative vomiting, in patients aged 18 years or older undergoing elective surgery under either TIVA or inhalational anaesthesia. The review inclusion criteria are presented in Box 1. Each title/abstract and full-text article were independently screened by two of four reviewers (HD, SM, JM, OG), and a third reviewer resolved conflicting decisions (MS, OG). One study published in Turkish was included in the analysis as the research team had the capacity to translate it into English[21] (Box 1).

#### Box 1. Systematic review inclusion criteria

**Study type**

Randomised controlled trials

**Language**

English or Turkish

**Population**

Adult patients undergoing elective surgery under either TIVA or inhalational anaesthesia

**Intervention**

Preoperative oral carbohydrate loading

**Control**

Overnight fasting

**Outcome**

Postoperative nausea and/or vomiting

Postoperative pain

Hospital length of stay

### Data extraction

Two of four reviewers (OG, NG, HD, LB) independently extracted data using a piloted Microsoft Excel spreadsheet. Two of four reviewers (NG, OG, LB, HD) assessed the risk of bias independently using the Cochrane ROB-2 Tool [22]. Conflicts were resolved by team consensus or were resolved by a third independent reviewer (MS). The corresponding authors of studies that met the inclusion criteria but did not report on the anaesthesia method were contacted. Those who responded to the inquiry of the type of anaesthesia were included in this review.

### Data synthesis

Meta-analysis was performed in the software package Stata BE 18 (64-bit; StataCorp LLC.) using the inverse-variance method with random effects models where data were available on the intervention and outcomes that met our inclusion criteria. Treatment effects for risk of PONV were expressed as risk ratio (RR). Standardised mean difference (SMD) was used for PONV severity measured using different scales and was interpreted according to Hedges’ g (0.2=small, 0.5=moderate, and 0.8=large) [23]. Hospital length of stay was expressed as weighted mean difference. Pain was measured using the Visual Analogue Scale (VAS). Studies were stratified into subgroups by the technique for general anaesthesia delivery (TIVA or inhalational anaesthesia). Heterogeneity was quantified using the I-squared statistic (I^2^), and values greater than 50% were considered to indicate substantial heterogeneity [24].

Studies that reported the risk of PONV without clear identification of either nausea or vomiting as separate events were analysed as combined PONV. Studies that reported medians and ranges were converted into mean and standard deviation (SD) [25]. Log risk ratio effect sizes were used to assess the risk of postoperative nausea and vomiting, as a combined or separate measure. We selected the period with the highest incidence or severity where outcomes were reported multiple times within the postoperative 24-hour periods. When studies reported postoperative vomiting or nausea occurrence separately, we considered post-operative vomiting results for meta-analysis, with the assumption that a patient with vomiting would also experience nausea. Two studies reported only postoperative nausea were also included as postoperative nausea and vomiting occurrence [26, 27]. In one study, SD was calculated from the reported standard error difference between the groups [28, 29]

We excluded a study from the meta-analysis due to limited outcome reporting, as the mean severity or incidence of postoperative PONV or pain could not be derived from available data [30]. We excluded studies where the anaesthesia type was not indicated, and the authors did not respond to our request for further information. A more detailed explanation for data synthesis can be found in **Appendix 1: Data Manipulations.**

## Results

### Study characteristics

Out of the 26 studies included in the review, one did not report the extractable data for outcomes (Figure 1). Therefore, 25 studies were included in the meta-analysis, which enrolled a total of 2,491 patients. Of those, 1,624 received inhalational anaesthesia (818 control; 806 intervention) and 867 received TIVA (435 control; 432 intervention) (Table 1).

**Figure 1.**
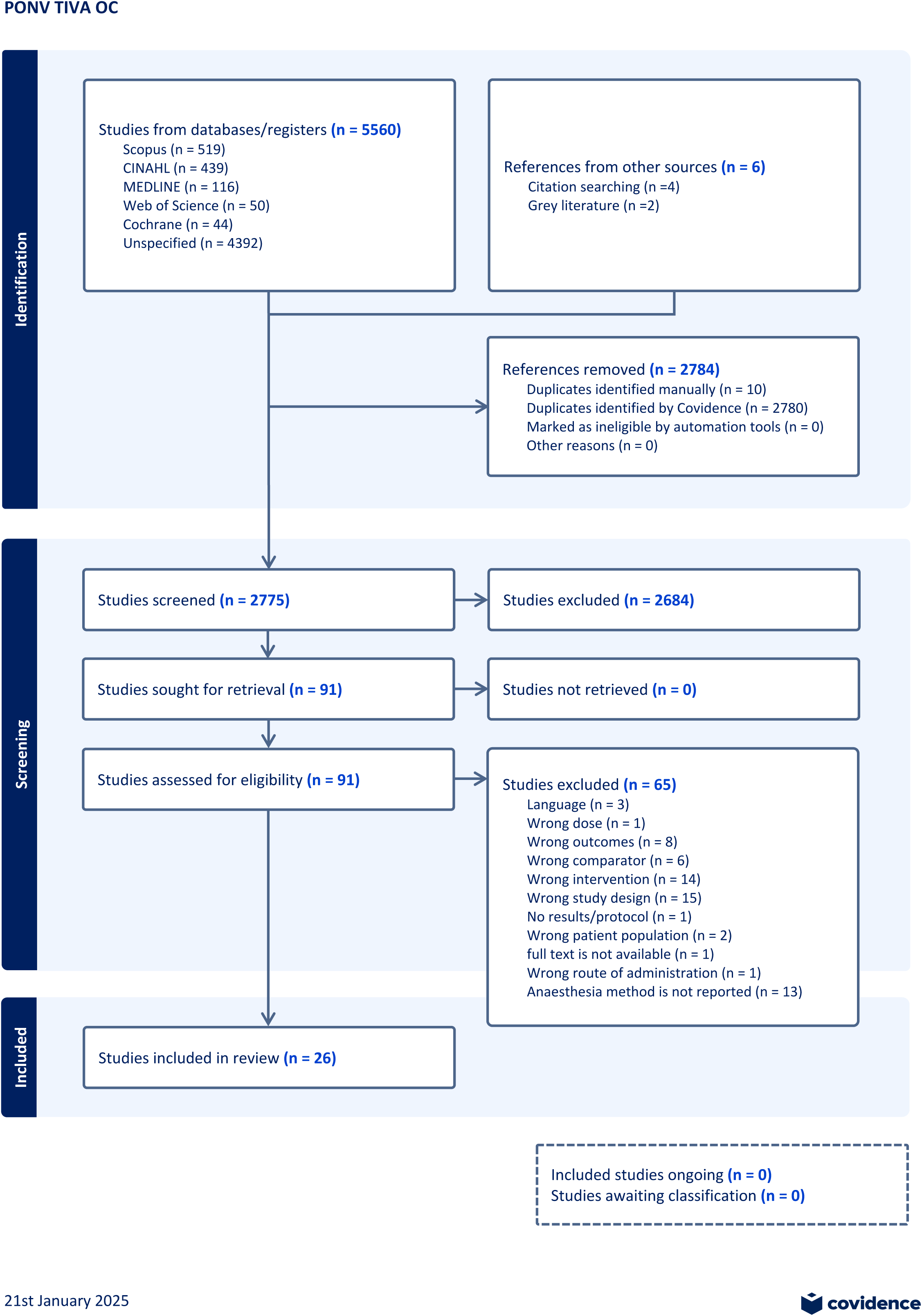
PRISMA Flow Chart.

**Table 1.**
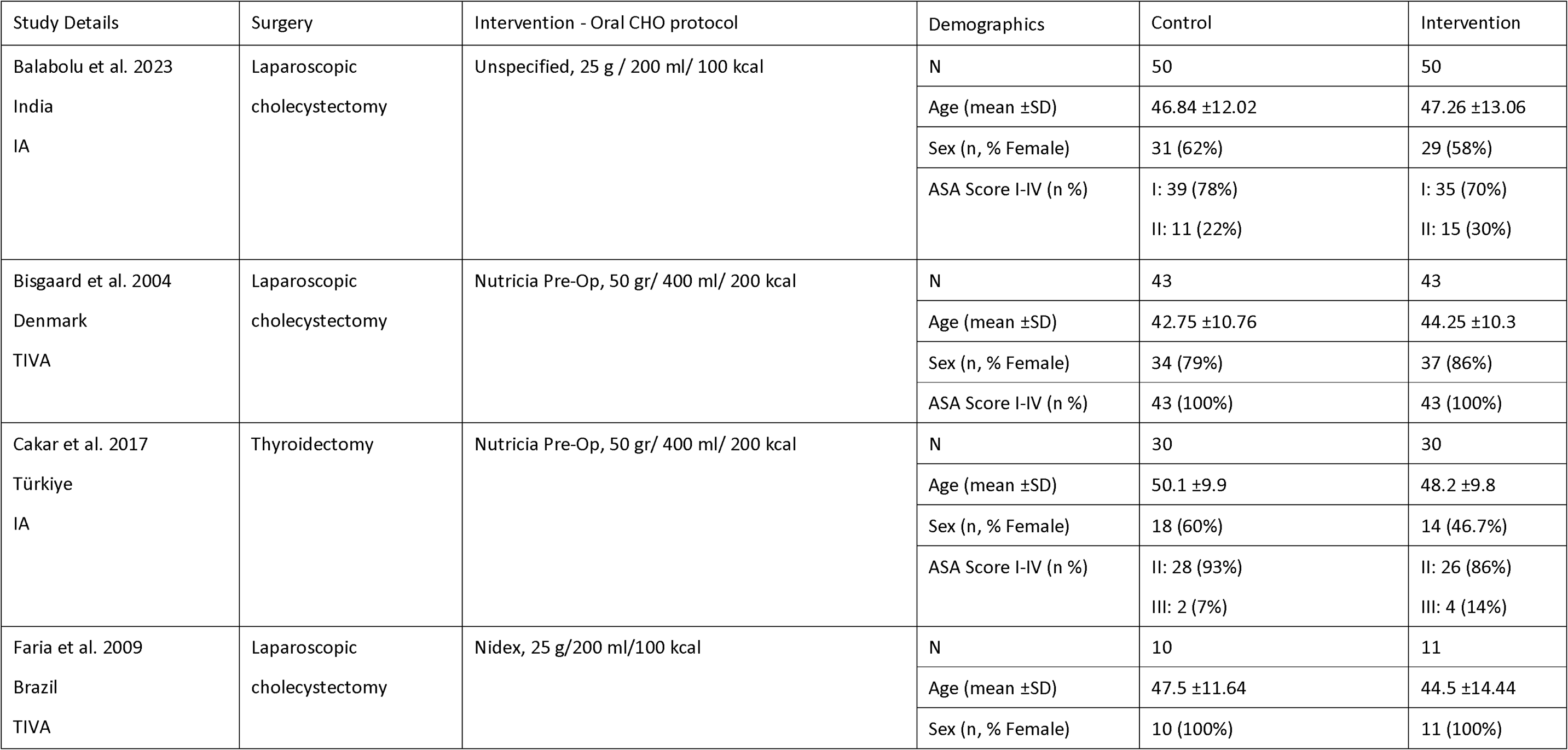
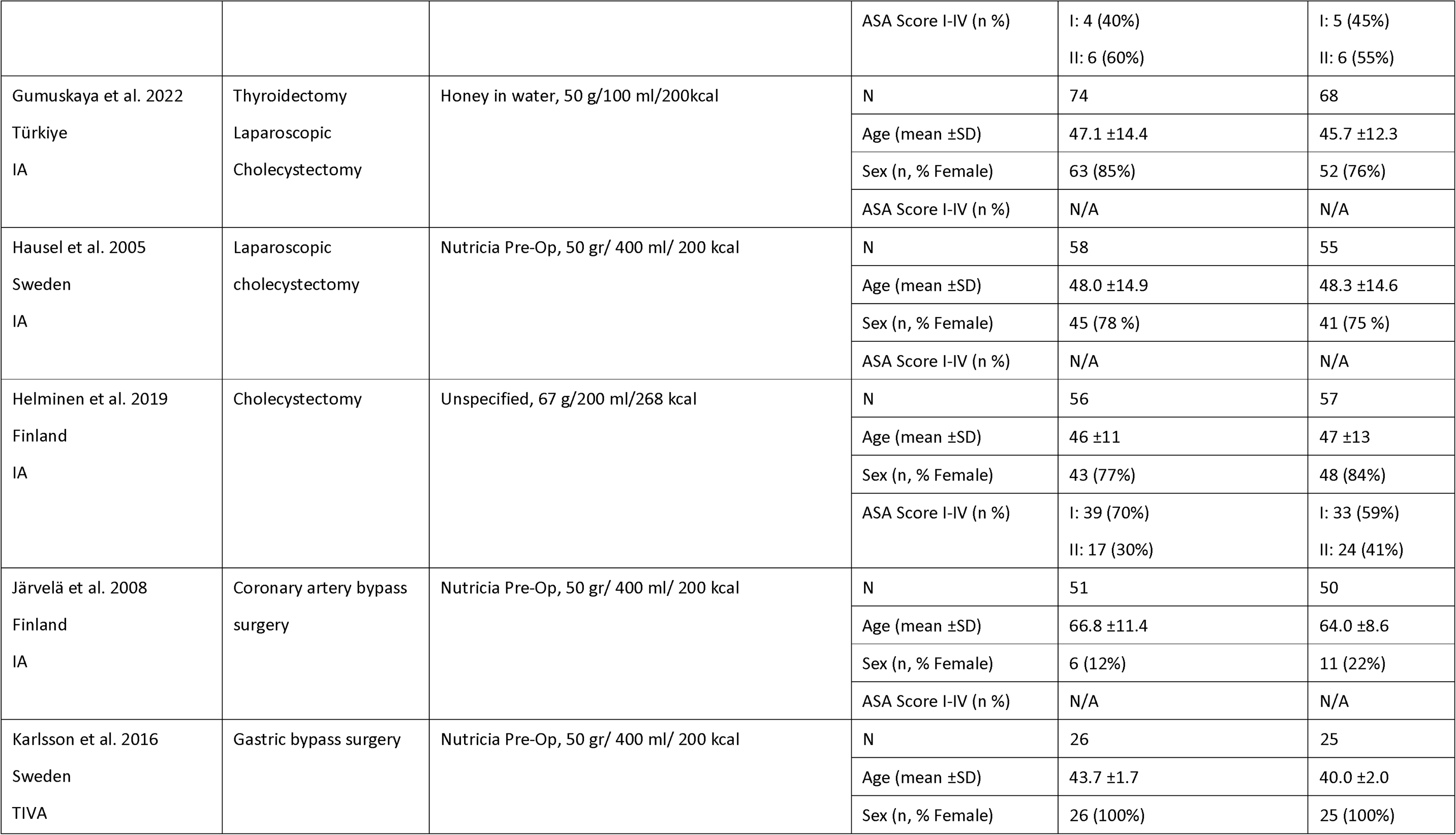
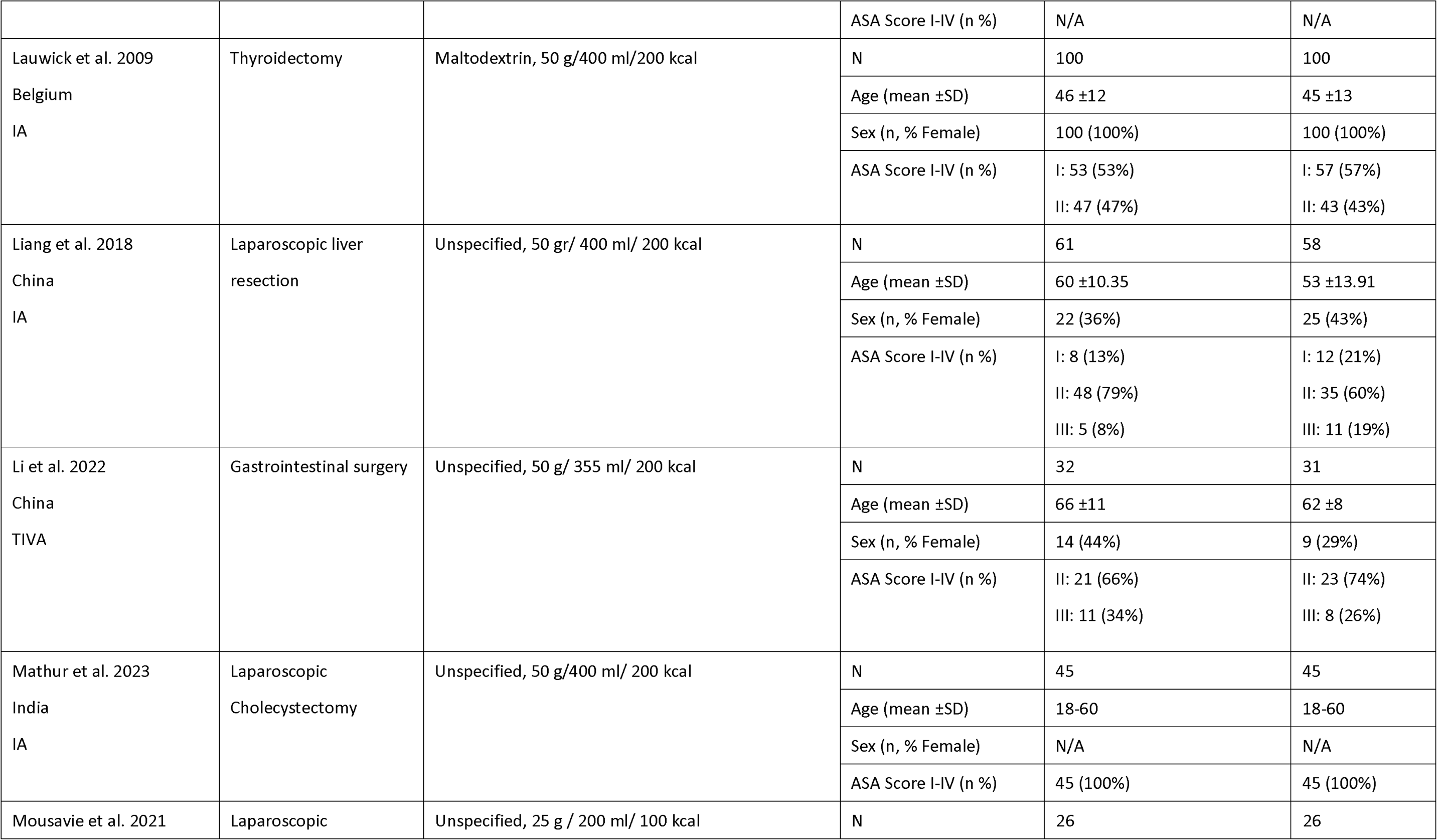
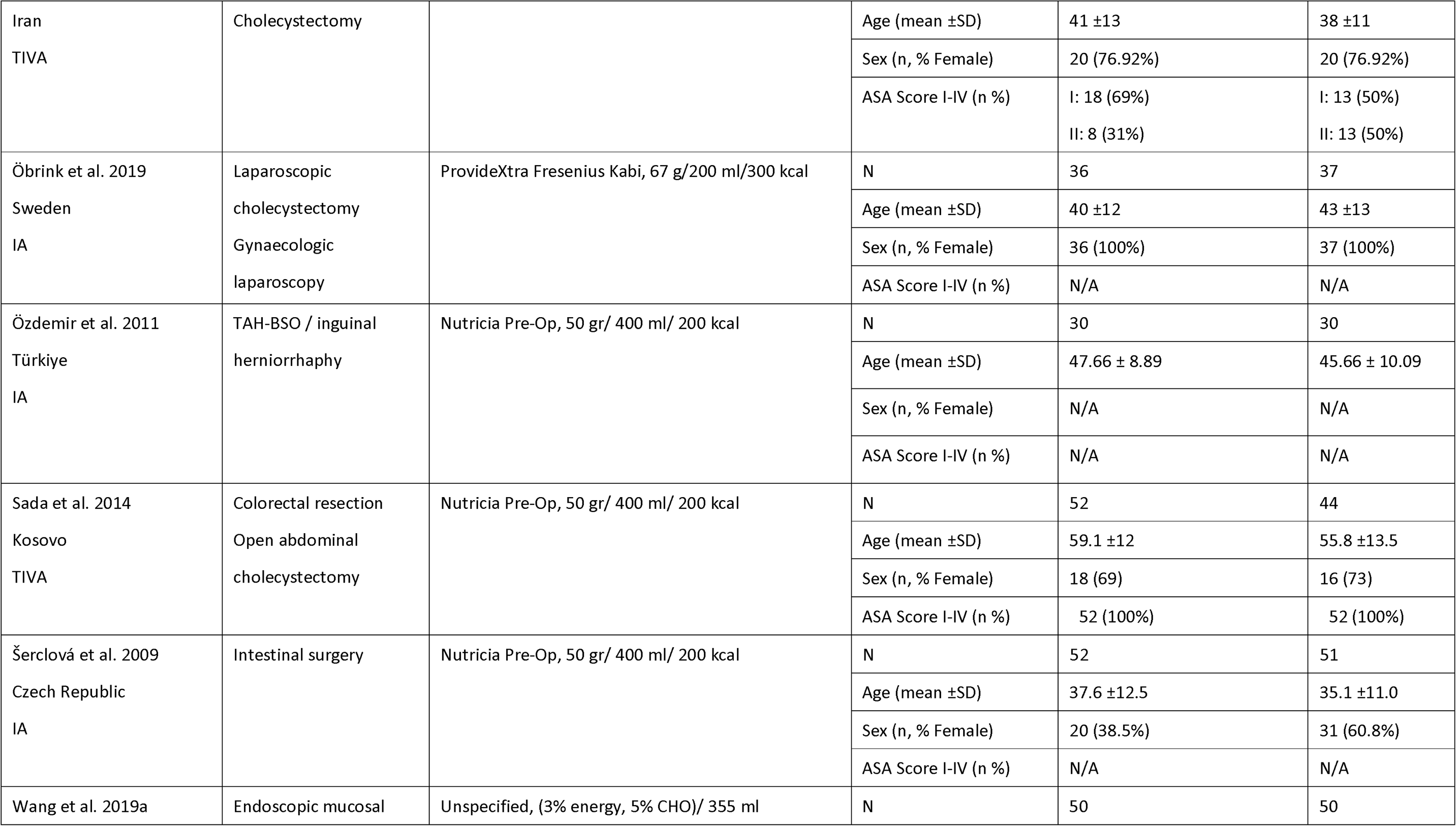
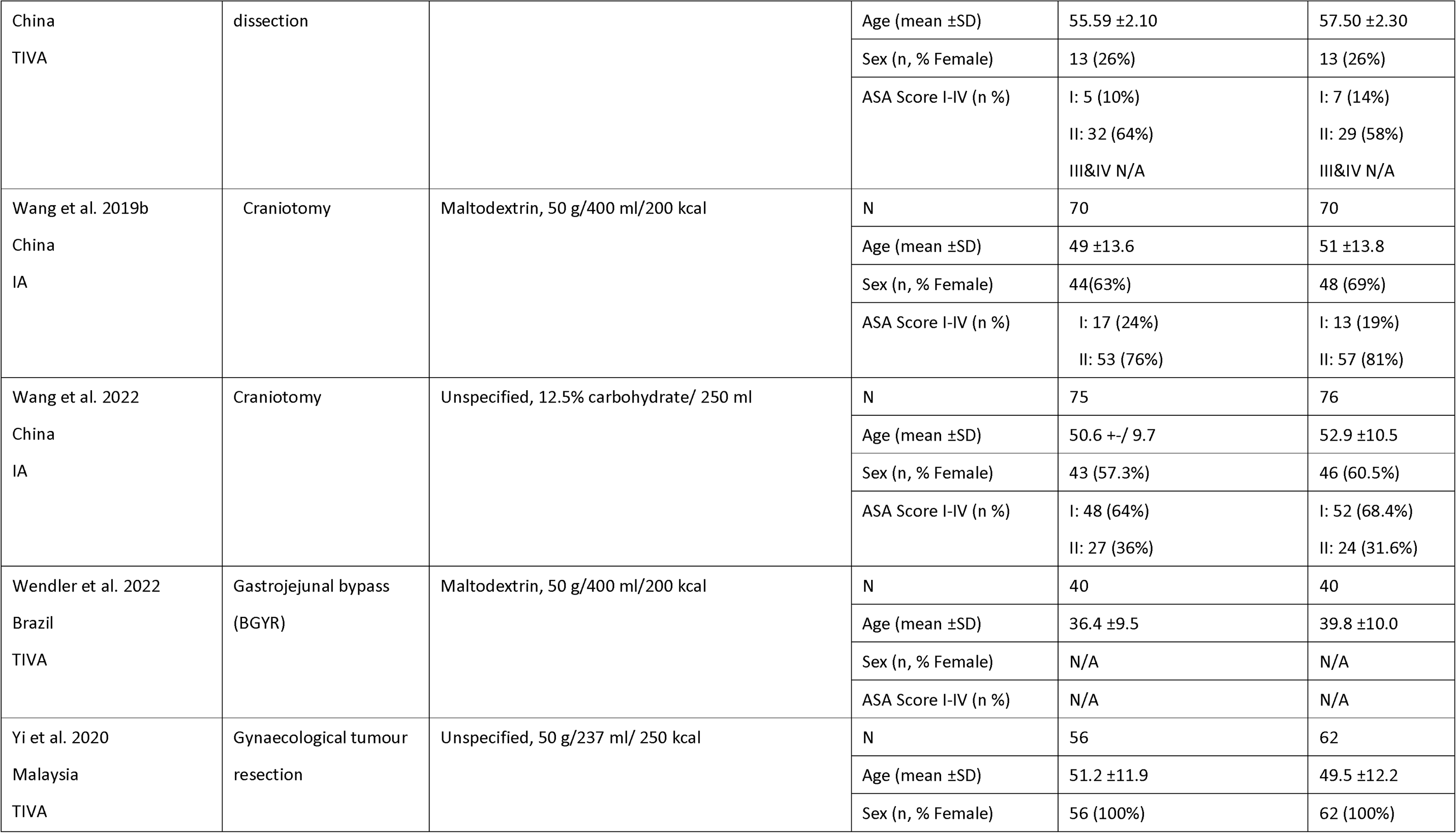
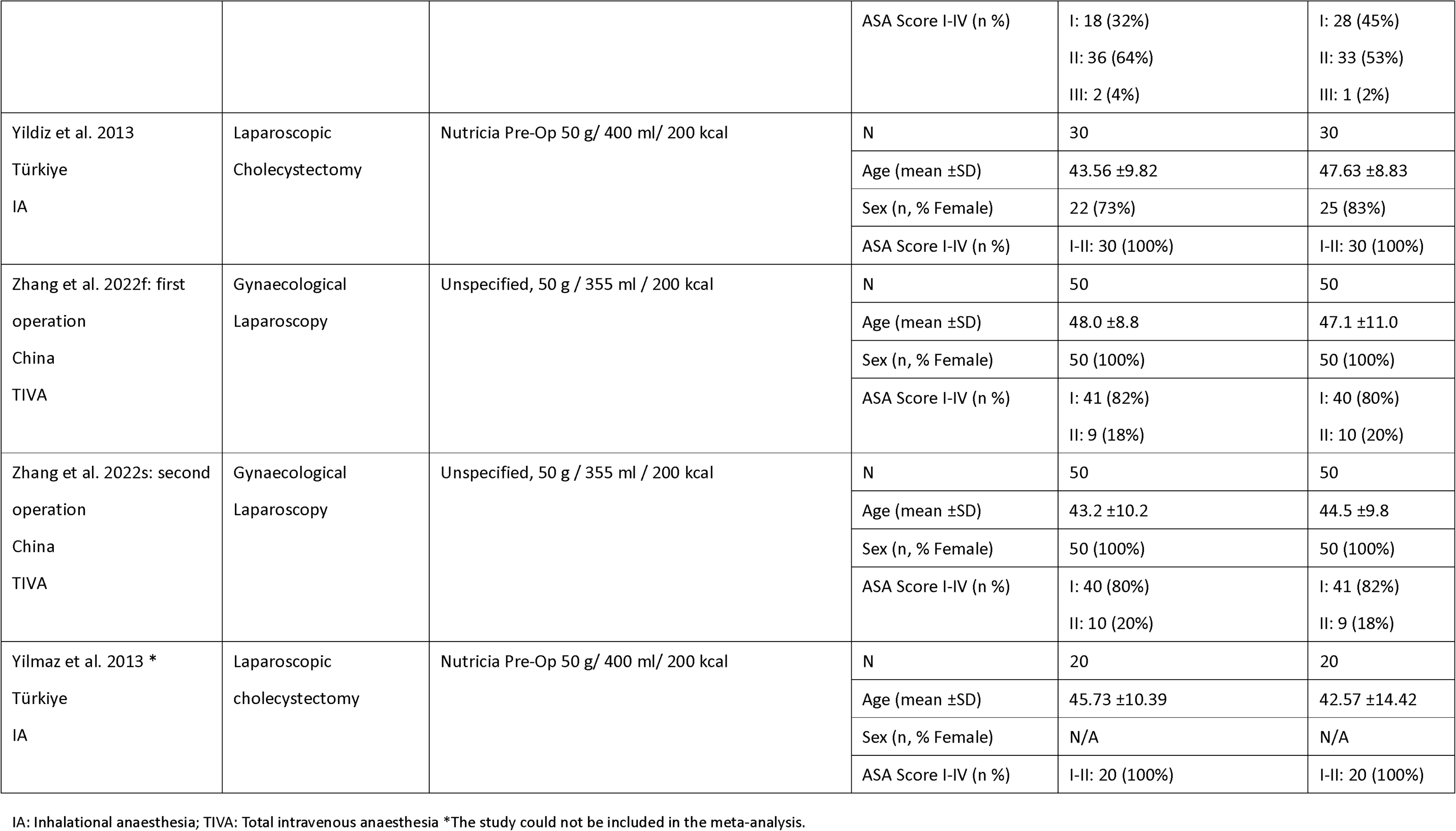
Characteristics of studies included in the review.

The studies were conducted in 12 countries: six from China, five from Turkey, three from Sweden, two each from Brazil, Finland, and India, and one each from Belgium, the Czech Republic, Denmark, USA, Iran, Kosovo, and Malaysia. Among the participants, 1,500 were female, and 656 were male (69% female; 31% male); four studies did not report gender data [21, 27, 30, 31]. The average participant age was 48 years, calculated by averaging the reported ages in the studies (Table 1).

The 16 studies reported PONV as binary data (count of occurrence), nine used either the Numeric Rating Scale (NRS) or Visual Analog Scale (VAS), and one used the Rhodes Index of Nausea and Vomiting (R-INVR) (Appendix 2: Data manipulations). For the secondary outcomes, 14 studies reported pain with VAS and 12 reported hospital length of stay.

### Risk of bias assessment

Risk of bias in the included studies is presented in Figure 2 [22, 32]. Out of 26 studies, 25 reported adequate randomisation methods and intended treatment. Two studies reported missing outcome data, two studies were at high risk for measurement of the outcome data, and four studies were at high risk for selecting the outcome results. Overall, six studies were evaluated to be at low risk, 15 demonstrated some concerns, and seven studies were at high risk for quality appraisal. No other potential sources of bias were identified for any of the included studies.

**Figure 2.**
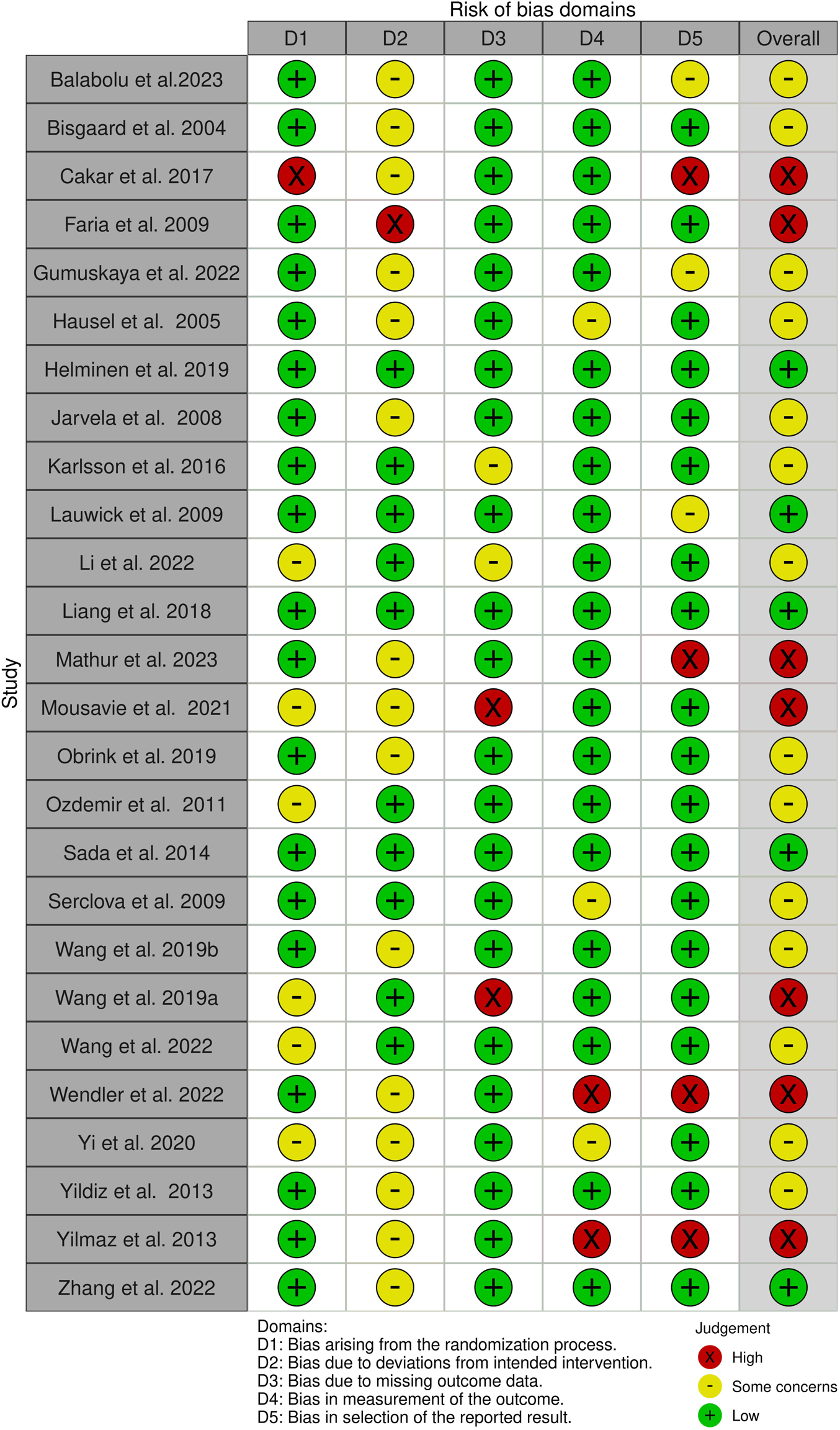
Risk of Bias.

### Severity of postoperative nausea and vomiting

The 10 studies measured the severity of PONV using a scale (VAS, NRS, or R-INVR). The overall pooled effect from 904 patients showed oral carbohydrate loading reduced PONV severity compared to overnight fasting, regardless of the type of anaesthesia (SMD: -0.46, 95% CI: -0.68 to -0.24, I^2^=62.09%). The impact appeared to be larger for TIVA patients (SMD: -0.54, 95% CI: -0.92 to -0.16, I^2^=46.18%) compared with inhalational (SMD: -0.43, 95% CI: -0.72 to -0.14, I^2^=72.24%) (Figure 3).

**Figure 3.**
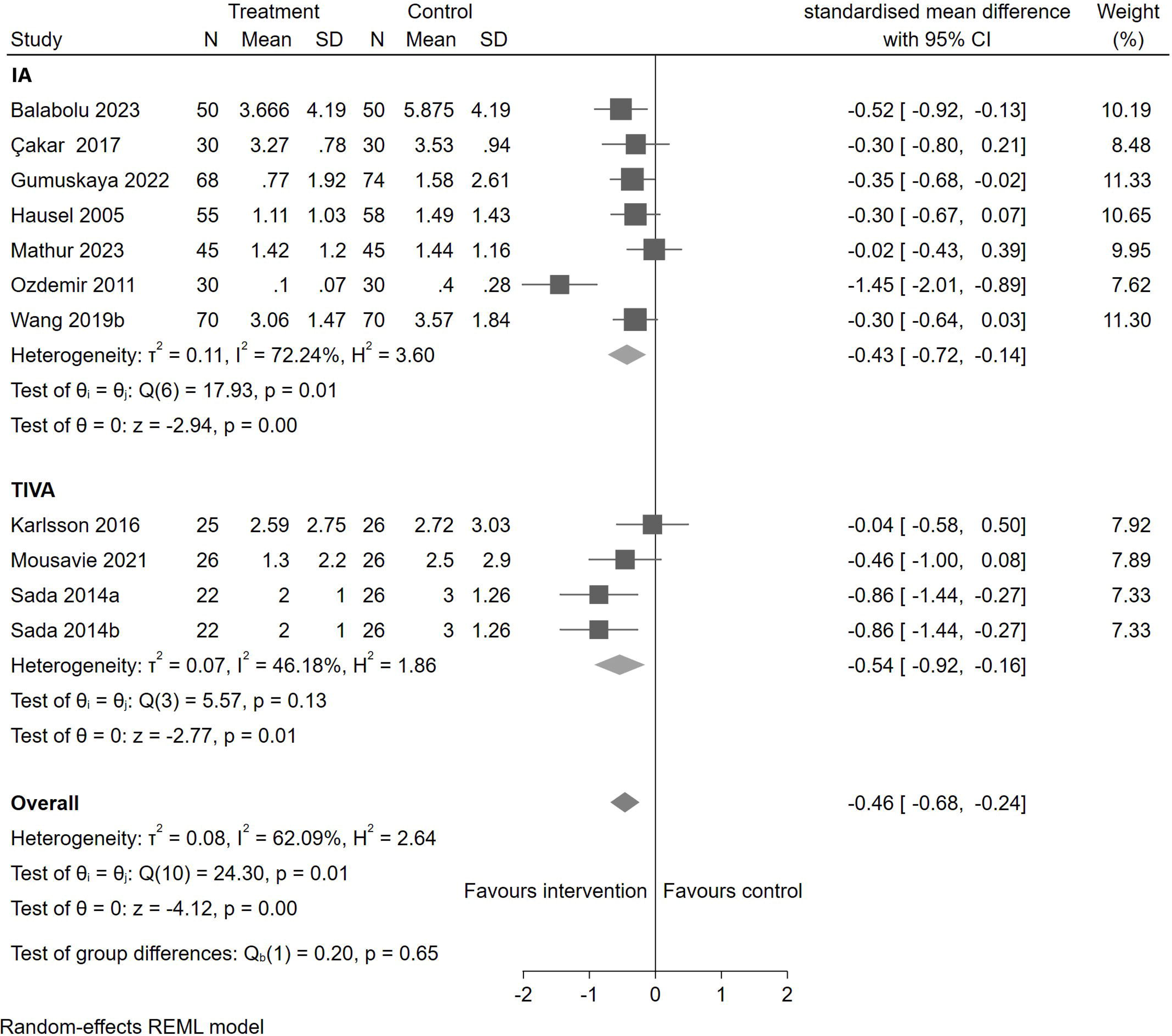
Postoperative nausea and vomiting severity with preoperative oral carbohydrate loading by anaesthesia approach (N = 904)

### Risk of postoperative nausea and vomiting

Of the 16 studies that reported on PONV risk as a binary outcome, the overall pooled effect from 1,774 patients favoured reduced PONV risk with preoperative oral carbohydrate loading (log risk ratio: –0.40, 95% CI –0.62 to –0.05, I^2^=53.70%) compared to overnight fasting when pooled across both anaesthesia approaches (Figure 4). When stratified by anaesthesia type, the effect was significant for adults receiving inhalational anaesthesia (log risk ratio: –0.26, 95% CI –0.52 to –0.01, I^2^=46.22%), but not for TIVA (log risk ratio: –0.40, 95% CI –1.02 to 0.22, I^2^=56.87%) (Figure 4).

**Figure 4.**
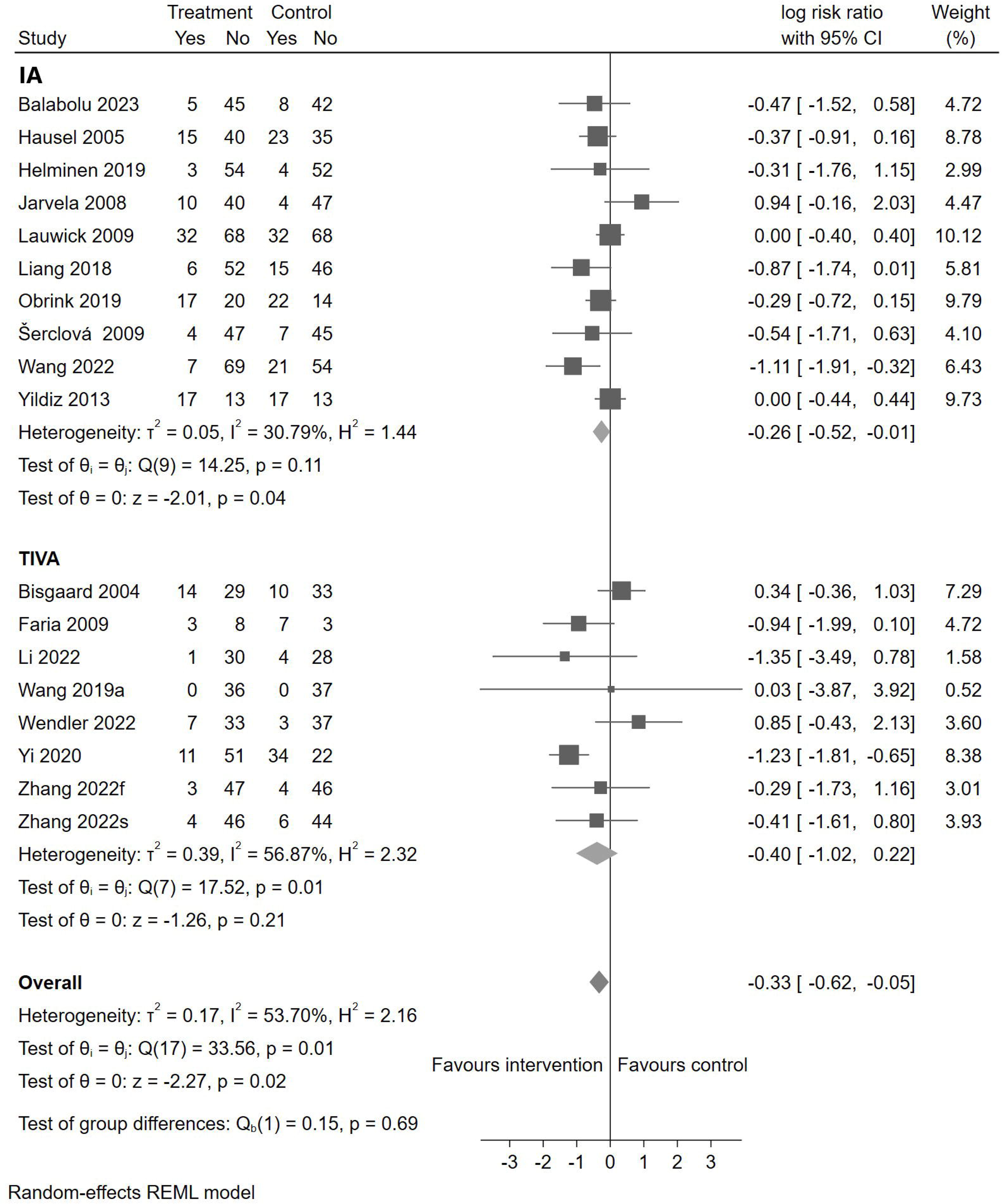
Postoperative nausea and vomiting risk with preoperative oral carbohydrate loading by anaesthesia approach (N = 1,774)

### Postoperative pain

In the 14 studies that reported on postoperative pain, the overall pooled effect of oral carbohydrate loading on 1,520 patients reduced postoperative pain severity (mean difference: -0.69, 95% CI: -1.13 to -0.25, I2=83.98%), compared to overnight fasting. The impact was significant for inhalational anaesthesia (mean difference: -0.79, 95% CI: -1.29 to -0.29, I^2^=85.59%) but not for TIVA (mean difference: -0.22, 95% CI: -0.91 to 0.46, I^2^=9.81%) (Figure 5).

**Figure 5.**
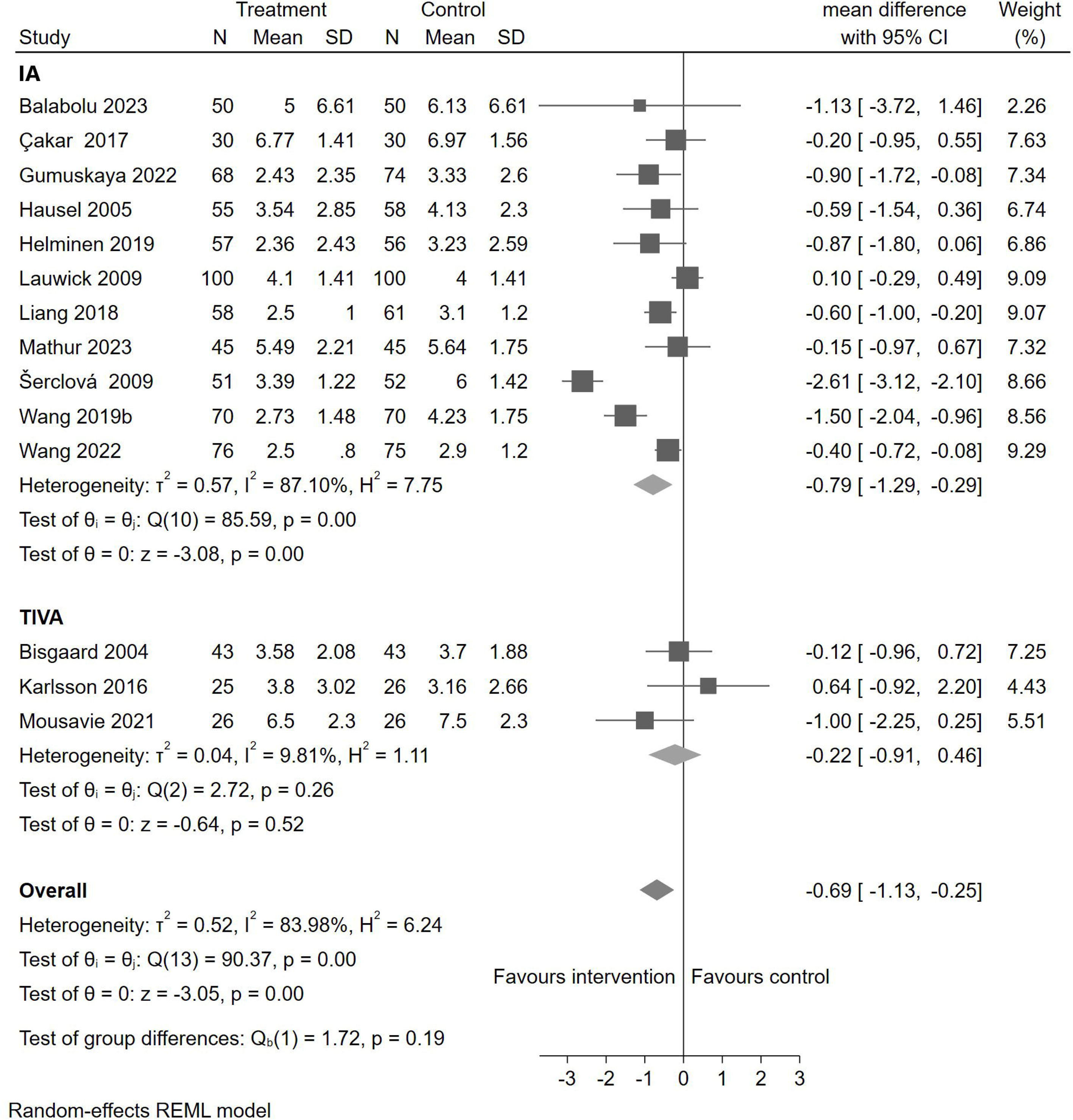
Postoperative pain levels with preoperative oral carbohydrate loading by anaesthesia approach (N = 1,520)

### Hospital length of stay

Twelve studies reported on hospital length of stay. The overall pooled effect from 1,310 patients showed oral carbohydrate loading did not impact hospital length of stay (mean difference: -0.18, 95% CI: -1.07 to 0.70, I^2^=99.61%), regardless of anaesthetic approach, compared to overnight fasting (Figure 6).

**Figure 6.**
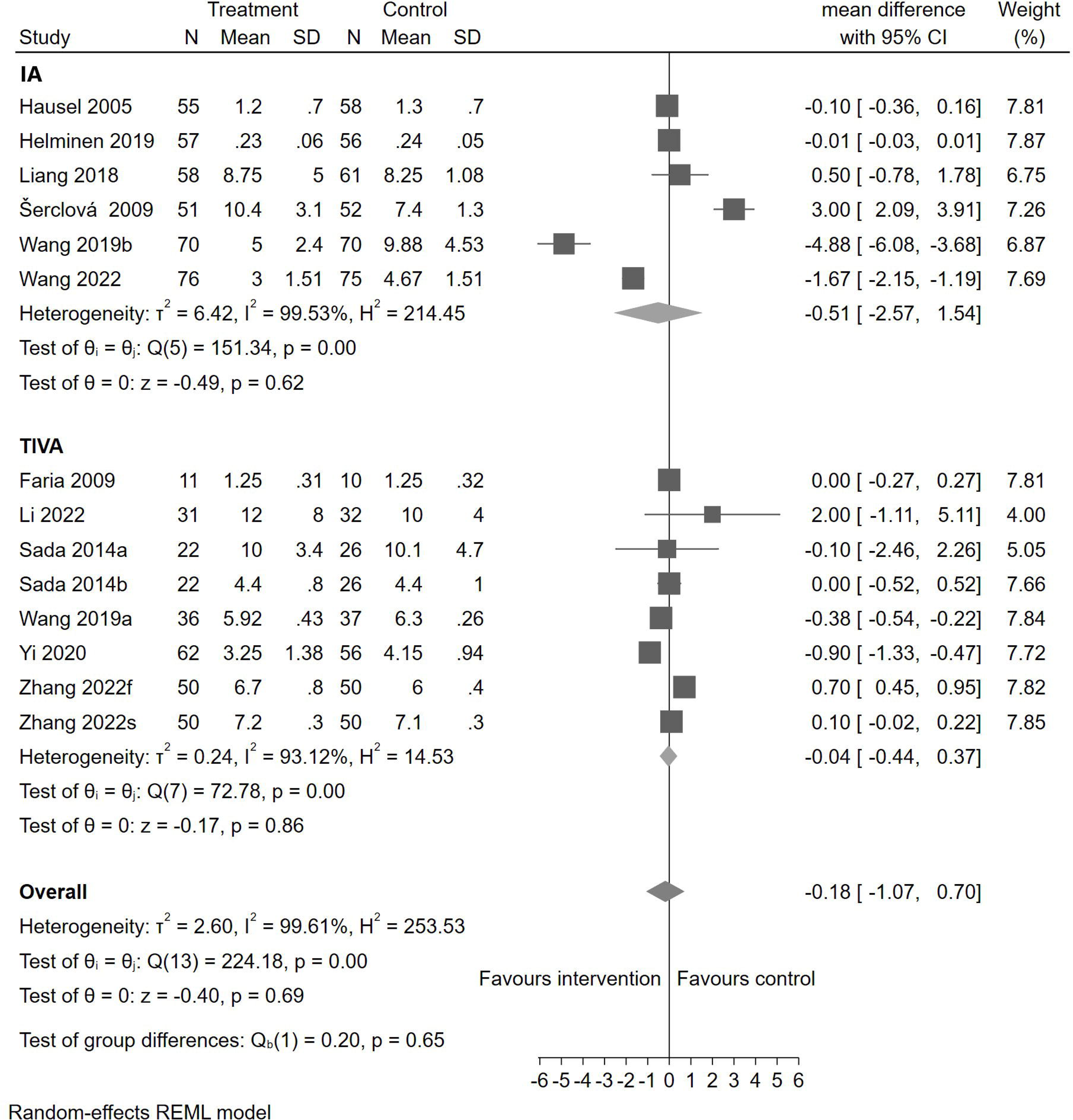
Postoperative length of stay with preoperative oral carbohydrate loading by anaesthesia approach (N = 1,310)

## Discussion

### Summary of findings

This systematic review and meta-analysis of randomised controlled trials demonstrated that oral carbohydrate loading significantly reduced the risk and severity of PONV when pooled across both anaesthesia approaches. The reduction in severity of PONV was more significant among TIVA patients. However, preoperative oral carbohydrate loading did not show a significant effect for the risk of PONV in patients receiving TIVA. Oral carbohydrate loading was associated with reduced postoperative pain severity, primarily among patients receiving inhalational anaesthesia. There was no significant impact on hospital length of stay regardless of the anaesthetic approach. One study that was not included in the meta-analysis reported significant improvements in PONV with preoperative oral carbohydrate loading [26]. Our findings indicate a potential role for preoperative oral carbohydrate loading in reducing the risk and severity of PONV. This meta-analysis reconciles previously conflicting findings in the literature regarding the effectiveness of oral carbohydrate loading on reducing the incidence of PONV, clarifying the potential role of anaesthesia approach [33–36].[33–36] Previous reviews have not always fully accounted for the interactions between preoperative oral carbohydrate loading in the presence of different anaesthesia techniques [37, 38]. Oral carbohydrate loading is thought to modulate metabolic responses, maintain blood glucose stability, and reduce the inflammatory response [13, 39–41]. Oral carbohydrate loading might therefore provide increased benefit in the presence of inhalational anaesthesia, given that inhalational anaesthesia has been shown to have less desirable metabolic effects compared to TIVA [1, 37, 38]. [37, 38]

The risk reduction of PONV with preoperative oral carbohydrate loading in the presence of TIVA is potentially confounded by several factors. Evidence supporting the clinical benefits of TIVA may mean that any additional benefits from carbohydrate loading could be limited to reducing severity. Additionally, PONV severity might be a more significant for patient discomfort [42–44].

PONV is a multifaceted concept with many confounders to consider, which were not reported consistently across the studies included in this systematic review. The well-established risk factors for PONV are female gender, non-smoking status, history of PONV or motion sickness, duration of anaesthesia, and use of perioperative opioid analgesics [1, 45]. Multimodal antiemetic prophylaxis is recommended by the Enhanced Recovery After Surgery (ERAS) Society, and several studies that implemented oral carbohydrate loading were also implementing comprehensive ERAS protocols, without PONV risk assessment [46, 47]. In some of the ERAS studies, the effect of reduced preoperative fasting on PONV could not be demonstrated reliably due to the confounding effect of multimodal antiemetic prophylaxis for most patients [46–48]. Fasting time was also unclear or inconsistent among participants. Most studies reported overnight or a minimum of eight hours of solid food fasting; however, the clear fluid or water fasting time varied among the control groups in some included studies [1, 38].

Previous systematic reviews have been unable to determine whether preoperative oral carbohydrate loading reduces postoperative pain, while Kampman et al. 2024 indicated a meaningful reduction in postoperative pain for patients receiving TIVA [33–35, 38, 41]. Our meta-analysis is the first to identify reductions in pain severity with oral carbohydrate loading, across both anaesthesia approaches, and more significantly for IA. However, high levels of heterogeneity should be considered interpreting this. Perioperative pain management, opioid use, the type of surgery, and patients’ ASA scores varied across studies.

The absence of effect on hospital length of stay (LOS), regardless of anaesthesia approach, is in line with some literature, however, contrasts with two meta-analysis, although these studies also indicated the insufficiency of evidence for LOS [15, 33, 35, 36, 49]. This could be explained by the increase in rapid discharge from fast-tracked or day surgery protocols. Despite the complexity of measuring an impact on perioperative hospital LOS, interventions as simple and cheap as oral carbohydrate loading could still benefit patient experience and reduce the cost of care [15, 33, 35, 36, 49].

### Limitations

Our meta-analysis was limited by the heterogeneity observed in some outcomes, particularly for PONV, such as the timing, tools used in measuring PONV, and other relevant assessment factors like reporting only nausea or vomiting. This heterogeneity underscores the need for further research to standardise reduced preoperative fasting protocols, assessment methods and reporting of their impact. Additionally, one study was excluded from the meta-analysis due to variations in reporting [30], suggesting a need for improved consistency in outcome measurement and reporting in this field. PONV was measured according to incidence, rate, rate difference, and severity using VAS (some studies referred to as NRS) and the Rhodes Index of Nausea, Vomiting, and Retching. Additionally, the PONV risk factors in the included study cohorts were not reported consistently to demonstrate this in the evidence summary. In addition, many studies reporting the effects of oral carbohydrate loading on PONV did not report the type of anaesthesia and, therefore, could not be included in this systematic review.

### Conclusion

This systematic review and meta-analysis suggest that preoperative oral carbohydrate loading could be a beneficial as a low-cost intervention to reduce PONV and pain, regardless of the general anaesthesia approach. Standardising reduced preoperative fasting protocols and timing specific to surgical contexts may enhance its effectiveness and allow more consistent patient outcomes. Integrating carbohydrate loading into preoperative care protocols could be beneficial, but further research is necessary to refine its application and establish stronger evidence and broader clinical guidelines.

## Supporting information

Appendix 2 Data manipulations

Appendix 1 Search strategy

PRISMA Check list

## Data Availability

The template data collection forms; data extracted from included studies; data used for all analyses; analytic code; any other materials used in the review, will be provided upon request.

## Acknowledgments

There are no competing interests to declare by any of the authors. The protocol was registered with PROSPERO (CRD42021222171). This research was supported by the Australian College of Perioperative Nurses 2021 Research Grant and the Australian College of Perianaesthesia Nurses 2023 Research Grant. We would like to acknowledge and thank Dr Sharon McGregor who assisted in abstract and title screening, and Dr David Rowe who assisted in conceptualising the study. Meta-analysis data is stored at the University of Sydney Research Data Management System and can be made available upon request. Mitchell Sarkies is supported by an NHMRC Investigator Grant (CIA Sarkies 2007970) and Sydney Horizon Fellowship. The template data collection forms; data extracted from included studies; data used for all analyses; analytic code; any other materials used in the review, will be provided upon request.

