## Appendix 2 Data manipulations for "The effectiveness of preoperative oral carbohydrate loading on postoperative nausea and vomiting in adults receiving total intravenous anaesthesia compared to inhalational anaesthesia: A systematic review and meta-analysis"

**Appendix 1 Data manipulations**

**PONV – binary**

Total of 856 patients’ PONV incidence were analysed [1-5]. Studies reported the incidence of postoperative nausea and vomiting (n= 856) without clear identification of either nausea or vomiting as separate outcomes were analysed as combined PONV incidence[2, 6, 7]. When early PONV (0-24 hours) was reported more than one period, we reported highest incidence period for analysis (6-24hr period) of to avoid erroneous overcounting [7].

**Nausea-binary**

Nausea incidences were analysed for 821 patients [6-13]. Studies reported nausea incidence within categories that were not well defined (ie., mild, moderate, and severe) also were analysed as incidence rate [7, 8]. Yildiz 2013 reported VAS scores for nausea which was converted to incidence, as means, medians or SD were not reported. The authors were contacted for comment and did not respond. Wendler 2022 reported nausea incidence as a percentage which were converted into whole numbers and rounded.

**Vomiting -binary**

Vomiting as an incidence were analysed for 991 patients [6-9, 12, 14-17]. Cakar’s response to a request for additional information allowed vomiting incidence. Wang 2019a reported both incidence of vomiting and PONV VAS scores as 0, including mean and standard deviation; therefore, was excluded from the analysis.

**PONV - Scales**

We used the 804 scale results [6, 15, 18-25]; standardised mean differences (Hedges g) were calculated due to the heterogeneity of the reported scales. Corresponding authors were contacted for information when data was not reported as mean and standard deviation. Cakar 2017 responded to our information request with raw data reporting VAS instead of the incidence rate ratio reported in their publication. The raw data used to calculate mean and standard deviation from Gumuskaya 2022 study; was merged (published data is separated by surgery type). Additionally, their inclusion of the R-INVR PONV score was not reported in this meta-analysis as they also reported data VAS scores for the cohort. Hausel 2005 and Serclova 2009 were estimated from graphical information using a ruler and paper. Hausel 2005, Helminen 2019, and Sada 2014 were estimated from median, range, and interquartile range [26].

Karlsson 2016 responded to our request for information and provided raw data which included means and standard deviations. Wang 2019b et al. reported PONV incidence in categories (1-4, 5-6, 7-10); we estimated the mean and standard deviation [26].

**Pain**

Pain was reported as VAS scores for 1420 patients [4, 6-8, 15, 16, 18-21, 25, 27]; if multiple scores were reported for VAS across 0-24hrs postop, the highest VAS score was considered for analysis[16]. Cakar 2017: responded to our information request with raw data reporting VAS instead of the incidence rate ratio. Gumuskaya et al. ‘s raw data were merged and used to calculate mean and standard deviation (separated by surgery type in their publication). Karlsson et al. reported headache and abdominal pain; as the surgery was abdominal, abdominal pain scores were used for analysis, received from the authors via email correspondence.

Lauwick 2009 and Serclova 2009 were estimated from mean and SEM to mean and SD. Bisgaard 2004, Hausel 2005, and Lauwick 2009 were converted from a scale of 0-100 to 0-10. Hausel 2005 and Serclova 2009 data were estimated from graphical information using a ruler and paper. Bisgaard 2004, Hausel 2005, and Helminen 2019’ s data were converted from median and range, and interquartile range [26]. Wang 2019b: the authors reported pain as incidence of people in separate categories (1-4, 5-6, 7-10) and we estimated the mean and standard deviation [26].

**Length of Stay**

The LOS was reported as days for a total of 1310 patients [4-6, 10, 12, 14-17, 24, 25, 27, 28] except Helminen 2019 and Yi 2020; which were converted from hours to days. Three of the twelve studies reported overall hospital LOS versus postoperative LOS; we combined this data with postoperative LOS as the patients were admitted to the hospital on the day of the surgery. However, Wang 2019b reported LOS from the end of the procedure as well as from admission; therefore, we analysed the latter for compatibility. The data from Faria 2009, Li 2022, Liang 2018, Wang 2109b, and Wang 2022 were estimated from median, range, and interquartile range to mean and standard deviation [26].

**Study characteristics**

We summarised evidence from 29 studies out of 5560 identified titles after 2784 duplicates were removed, and 91 were considered potentially eligible for inclusion criteria and received full-text review (Figure 1). In addition to the meta-analysis conducted with 24 studies; we also included additional five studies that were not appropriate for meta-analysis in the evidence summary table to demonstrate the significance of PONV outcomes, demonstrate the confounders and risk factors that impact PONV, pain, and LOS.

There were no major apparent differences between intervention and control groups regarding PONV risk factors of age, gender, history of PONV and motion sickness, the duration of anaesthesia, the use of antiemetics and opioid painkillers among the studies reported this data (Table 1).
