## Appendix 1 Search strategy for "The effectiveness of preoperative oral carbohydrate loading on postoperative nausea and vomiting in adults receiving total intravenous anaesthesia compared to inhalational anaesthesia: A systematic review and meta-analysis"

**Review questions:**

Does oral carbohydrate administration reduce PONV incidence and severity for adult patients undergoing elective surgery receiving total intravenous anaesthesia compared to overnight fasting?

Does total intravenous anaesthesia reduce the risk of PONV incidence and severity compared to inhaler and intravenous anaesthesia; for adult patients undergoing elective surgery?

**Search Strategy:**

Pub-Med/MEDLINE, Cochrane, CINAHL, Elsevier/Scopus, ScienceDirect, Wiley, Clinical Key, and Google Scholar

**Boolean Search Key words:**

“postoperative nausea and vomiting” OR “PONV” OR “ponv”

AND

“operating room” OR “operating theatre” OR “perioperative” OR “preoperative” OR “postoperative” OR “perianesthesia” OR “perianaesthesia” OR “postanesthesia” OR “postanaesthesia” OR “volatile anaesthesia” OR “inhaler anesthesia” OR “inhalation anesthesia”OR “total intravenous anesthesia” OR “intravenous anesthesia”

AND

“carbohydrate” OR “loading” OR “calorie” OR “fasting” OR “glucose”

**Search query:**  (“nausea” OR “vomiting” OR “PONV” OR “ponv”) AND (“surg*” OR “operat*” OR “peri?op*” OR “pre?op*” OR “post?op*” OR “perianest*” OR “peri?anaest*” OR “post?an*” OR “volatile an*” OR “inh*” OR “anaest*” OR “anest*” OR “total intravenous an*” OR “TIVA”) AND (“carbohydrate” OR “loading” OR “calorie” OR “fasting” OR “glucose”)
